# Pandemic Policy Design Via Feedback: A Modelling Study

**DOI:** 10.1101/2021.09.23.21263924

**Authors:** Klaske Van Heusden, Greg E. Stewart, Sarah P. Otto, Guy A. Dumont

## Abstract

**Background:** The COVID-19 pandemic has had an enormous toll on human health and well-being and led to major social and economic disruptions. Public health interventions in response to burgeoning case numbers and hospitalizations have repeatedly bent down the epidemic curve in many jurisdictions, effectively creating a closed-loop dynamic system. We aim to formalize and illustrate how to incorporate principles of feedback control into pandemic projections and decision making.

**Methods:** Starting with a SEEIQR epidemiological model, we illustrate how feedback control can be incorporated into pandemic management using a simple design (proportional-integral or PI control), which couples recent changes in case numbers or hospital occupancy with explicit policy restrictions. We then analyse a closed-loop system between the SEEIQR model and the designed feedback controller to illustrate the potential benefits of pandemic policy design that incorporates feedback.

**Findings:** We first explored a feedback design that responded to hospital measured infections , demonstrating robust ability to control a pandemic despite simulating large uncertainty in reproduction number *R*_0_ (range: 1.04-5.18) and average time to hospital admission (range: 4-28 days). The second design compared responding to hospital occupancy to responding to case counts, showing that shorter delays reduced both the cumulative case count and the average level of interventions. Finally, we show that feedback is robust to changing public compliance to public health directives and to systemic changes associated with new variants of concern and with the introduction of a vaccination program.

**Interpretation:** The negative impact of a pandemic on human health and societal disruption can be reduced by coupling models of disease propagation with models of the decision-making process. This creates a closed-loop system that better represents the coupled dynamics of a disease and public health responses. Importantly, we show that feedback control is robust to delays in both measurements and responses and to uncertainty in model parameters and the efficacy of control measures.

**Funding:** A Natural Sciences and Engineering Research Council Grant to SPO (RGPIN-2016-03711). NSERC Discovery Grant to GAD (RGPIN-2018-05151).

Research in context

Evidence before this study
A search on Pubmed on August 24 2021 for the keywords ((COVID[Title/Abstract]) AND (model[Title/Abstract])) AND ((projection[Title/Abstract]) OR (forecast[Title/Abstract])) returned 422 articles, reflecting challenges in accurate modeling of the pandemic. This pandemic has put both the influence and limitations of modeling into the public eye. A google search with the keywords “covid death model” AND “wrong” returns scientific articles as well as opinion and newspaper articles including in influential media. Many of the modeling papers include simulation scenarios evaluating possible interventions and control strategies. We found very few articles that take the inherent feedback into account that happens when intervention policies are determined in practice. An exception is the widely simulated ad hoc on-off policy where interventions are triggered when case or hospitalization levels are reached. A google scholar search using the keywords “covid 19 feedback control” returns over 30 articles in engineering journals that explicitly address the inherent feedback in the decision-making process. Many of these articles are highly technical and one of our goals in this study is to connect the engineering community with the public health and epidemiology community.

Added value of this study
This study aims to provide the public health community with a brief overview of feedback control principles and how they apply to decision-making for a pandemic such as COVID-19. In this work we shift the focus from prediction to the design of interventions. We used a standard SEEIQR model with a simple controller to simulate the implication of feedback in the decision-making process. We show that, in contrast to highly varying open-loop projections, incorporating feedback explicitly in the decision-making process is more reflective of real-world situations and illustrates that effective decision making can be made even with only moderately accurate models. We show that effective feedback policy can be designed using daily case counts or hospitalizations and that it is not necessary to know the fraction of cases that is detected to control the pandemic. We illustrate how fundamental limitations of feedback impact the achievable level of control.

Implications of all the available evidence
It is recommended that models of propagation of the virus be augmented with models of the decision-making process to produce a closed-loop system that is more representative of the real-world situation. Policy decisions resulting from systematic intervention design, rather than ad hoc decisions, will make projections more reliable and can encourage taking earlier and smaller actions, which reduce both case counts and the severity of interventions. Using feedback principles, effective control strategies can be designed even if the pandemic characteristics remain highly uncertain. Case numbers as well as total interventions can be reduced by minimizing all delays in the chain of information from testing and reporting to decision making and public response.

## 1. Introduction

With over 220 million cases and 4.6 million deaths (WHO dashboard, accessed September 12, 2021), the COVID-19 pandemic has placed an enormous burden on individuals, health systems, and society. In order to reduce the rate of transmission of the disease, most countries have implemented similar measures, including lockdowns, closures of restaurants, bars and non-essential services, school closures, curfews, prohibition of large gatherings, obligatory mask-wearing, travel restrictions, testing and tracing, and more recently mass vaccination. All along, public health authorities and governments have had to adjust and modulate those public health measures in response to a very fluid and rapidly evolving dynamic situation. Deciding which intervention to implement and when remains challenging: i) epidemiological models predicting the course of the epidemic show significant uncertainty [1, 2, 3] ii) the efficacy of individual measures remains poorly known and may change over time [4] and iii) there is hesitancy to impose strict measures that have significant social, health and economic implications. In order to do so, decision-makers use up-to-date measures such as daily or weekly incidence rates, hospital admissions and ICU occupancy rate to set restrictions. They are thus implicitly using feedback.

So, just like Molière’s Mr Jourdain^1^ who “For more than forty years (has) been speaking prose without knowing anything about it”, public health officers and decision makers have been using feedback in an ad-hoc manner without knowing feedback theory or the full range of tools that it offers.

The aim of this paper is thus to provide the public health community a brief overview of basic feedback control principles and an illustration of how to use them in the decision-making process when designing interventions to manage a pandemic such as COVID-19. Pandemic control is, at its core, not just about disease spread but about behavioural and policy reactions to the disease (i.e., feedback). The vast literature on control theory is, however, largely unknown to epidemiologists and public health decision makers. In the context of the current COVID-19 pandemic, we highlight control principles that could help pandemic management, with the goal of reducing both case counts and social impacts of restrictions.

Explicit consideration of feedback shifts the focus from prediction to design of interventions. COVID-19 predictions show large uncertainty in part because they are scenario based, where one fixed intervention sequence is used to predict possible outcomes (open-loop). In reality, the decision-making process will assess outcomes and adjust accordingly; if the pandemic continues to grow, additional interventions are likely put in place. This decision-making process uses feedback, even if used informally, implicitly or inconsistently. By representing control of COVID-19 explicitly in a feedback framework, the impact of such reactive measures (closed-loop) can be systematically and rigorously analyzed. Instead of implementing ad-hoc policies, transparent and effective policies can be planned and optimized using feedback theory.

Feedback is a powerful tool when wielded carefully. It can permit aggressive decision-making under uncertain conditions. However when used inappropriately it can also introduce undesirable behaviors. Too conservative a design is safe but will respond too slowly to a changing environment and disturbances. An overly aggressive design is too sensitive to model uncertainty, which may result in wild swings in behaviour or exponential growth.

Implementing an appropriate feedback design can reduce the health, social, and economic costs and also be more robust as the pandemic changes over time, through viral evolution or ongoing vaccinations. Such unpredictable changes can be dealt with via consistent use of feedback based on measurements reflecting the state of the pandemic, even in the presence of significant and uncertain time delays.

Figure 1 illustrates the range of outcomes that can result from apparently subtle changes in interventions, applied to a COVID-19 epidemiological model. It also shows that small early interventions can avoid large interventions later. Hesitancy in implementing control measures risks a greater toll from the disease. The goal of this paper is to demonstrate how this sensitivity can be harnessed and used to advantage in designing aggressive yet robust public policies even under conditions of high uncertainty. While this will not make difficult decisions easier, it will make them more transparent, informed, and effective.

**Figure 1.**
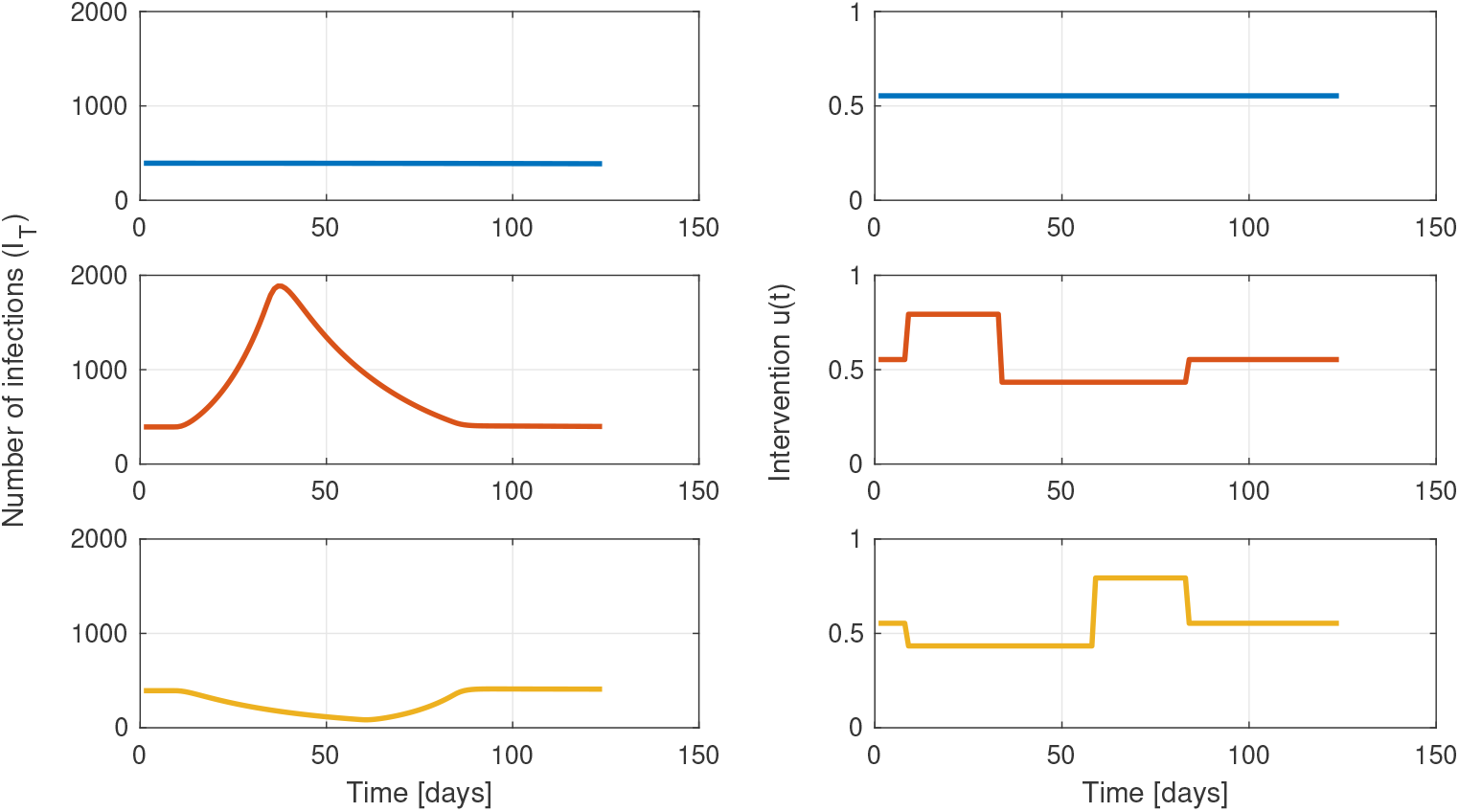
High sensitivity to interventions: The average level of interventions, as well as the initial and final daily case rates, are the same for all three scenarios. In the first scenario, interventions that lead to zero growth from the initial *I*_*T*_ = 400 are maintained, leading to a cumulative infection case count of ∫ *I*_*T*_ (*t*)*dt* of 48516. In the second scenario, restrictions are initially relaxed, leading to pandemic growth. Tighter restrictions are then implemented to return to the initial number of infections *I*_*T*_ , leading to almost twice the number of total infections: ∫ *I*_*T*_ (*t*)*dt* = 95174 illustrating the cost of delayed action. The third scenario starts by implementing stricter interventions followed by relaxation, resulting in ∫ *I*_*T*_ (*t*)*dt* = 34821, a reduction of 28% from the first case despite the same average level of restrictions. The variable *u*(*t*) represents the activity level of those who can socially distance, ranging from *u*(*t*) = 0 with complete distancing to *u*(*t*) = 1 for normal levels of activity (see Appendix A).

## 2. Methods

### 2.1. Models

Simulation results are based on a two-group compartmental SEEIQR model which was developed to estimate the effect of social distancing in early 2020 in British Columbia, Canada [5]. As the focus of this study is the decision-making rather than the modeling, only a brief overview of the simulation model follows, details may be found in the Appendix.

This model contains two groups, one representing individuals who are socially distancing and one representing individuals who cannot distance, for example due to their profession. The model compartments represent the number of susceptible (S), exposed (E1), pre-symptomatic and infectious (E2), symptomatic and infectious (I), quarantined (Q), recovered or deceased (R) for each group. The recovered (R) group does not affect infection rates and is omitted in this work. The model was extended to include vaccinations and the appearance of new variants as detailed in the Appendix.

The effect of social restrictions is introduced in this model through the variable *u*(*t*) in (7)-(9), which can vary with time and affects transmission rates involving the distanced group. Public health policies are assumed to impact *u*(*t*), where *u*(*t*) = 0 represents completely restricted social interactions with no transmission among distancing individuals and *u*(*t*) = 1 represents normal activity levels.

Next we define the variable to be measured to inform the policy decisions. While the model (7)-(9) contains four groups (unable or able to socially distance, non-variant or variant), any measure of infections or hospitalizations represents the total of these groups: *I*_*T*_ (*t*) = *I*(*t*) + *I*_*d*_(*t*) + *I*_*v*_(*t*) + *I*_*dv*_(*t*). The epidemiological model detailed in Appendix A describes how the number of cases in each infected group changes over time. The exact number of infected individuals, *I*_*T*_ (*t*), is not known but is estimated by some measure in a lagged fashion:

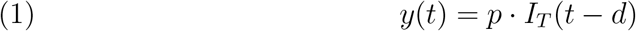

where *p* represents the fraction of the infected population being identified or measured and *d* represents the delay in days of before measurement. In this paper we will contrast two different measures. In Scenario 1 we seek to represent hospital occupancy, and we set *p* = 0.073 and a delay of *d* = 14 days.^2^

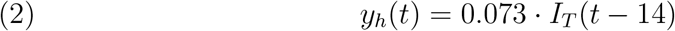

Scenario 2 is intended to reflect detecting new cases with fast public testing, and there we set a much shorter delay of *d* = 2 days, but an unknown fraction *p* of tests performed:

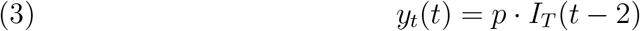

Note that the observed cases *y* will generally differ from the actual cases depending on the rates of false positive (FP) and false negative (FN) rates in the test used. We will discuss this point further in Section 2.2.

### 2.2. Systematic feedback design of interventions

There are many techniques developed, deployed, and industrially validated over several decades for the design of feedback control systems, see for example [6]. Generally, these techniques allow the designer to balance the speed of control against the harms of overreacting, especially given system noise, delays, and model uncertainty.

We will assume that pandemic intervention policies are defined by a function:

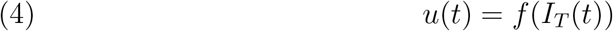

meaning that there is some rule, *f* (), that translates the number of infected cases *I*_*T*_ (*t*) into public health measures, *u*(*t*), that affect transmission among those individuals able to socially distance (see Appendix A). The formulation in (4) is general enough to cover most policies, whether ad hoc heuristics or more systematic approaches. The goal is then to design this function such that the resulting closed-loop delivers the desired performance and stability.

We select a straightforward control design technique known as SIMC (simple internal model control), originally developed for process control applications, where dynamics are often complex and nonlinear [7]. We selected SIMC and the particular type of controller it implements (“proportional-integral-derivative” PID) because they are more straightforward to describe and to implement, improving the chance of successful application of feedback systems for pandemic control.

The SIMC design method includes the following steps: 1) Obtain a linear first- or second-order plus time-delay model approximating the process. 2) Depending on the characteristics of the approximate model, the SIMC rules suggest a controller structure and controller parameters as a function of the estimated model parameters.

In this design technique we thus use a linear approximation of the nonlinear SEEIQR model to synthesize the controller, but all simulations of that control policy will be performed against the full nonlinear model in Appendix A. The linear approximation is obtained in three steps that are described in detail in the Appendix. First, the number of susceptible individuals is assumed constant, *S*(*t*) = *S*_0_. This assumption is commonly used to linearize epidemiological models. The relationship between the social activity level, *u*(*t*), and the measure of cases, *y*(*t*), remains nonlinear. Second we transform the measured cases variable *z*(*t*) = ln(*y*(*t*)) and the intervention variable *v*(*t*) = *u*(*t*) − *u*_0_, where *u*_0_ is the unknown level of restrictions required to achieve zero pandemic growth. This results in an approximate linear relationship^3^ in the dynamics relating transformed inputs *v*(*t*) and transformed states *z*(*t*) and is insensitive to scaling. Lastly, the transformed model is injected with a series of steps in intervention *u*(*t*) to obtain an input-output response over a range of operating points and a linear dynamic model^4^ fit to the response using a non-linear least squares estimation (the output-error method [8]):

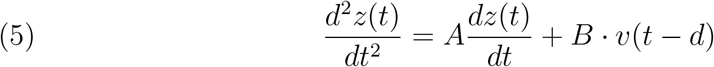

resulting in constants *A* = − 0.5787 and *B* = 0.1572 for the parameters of model (7)-(9), as described in Appendix C.

Next, for systems such as (5) where the response delay, *d*, is significant, the SIMC method [7] recommends a proportional-integral control policy:

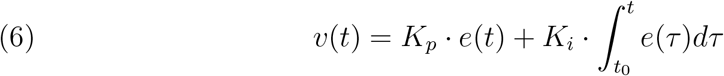

where *e*(*t*) = *r*(*t*) − *z*(*t*) is the error between the feedback values *z*(*t*) and their desired value, setpoint *r*(*t*) (which in practice would typically be set to correspond to a number of cases that is smaller than the healthcare capacity of the region). The format of this controller is the structure used in over 90% of control engineering applications [9]. The integrator in the second term in feedback controller (6) has the desired effect of driving measured outputs to the setpoint *z*(*t*) = *r*(*t*) in spite of model uncertainty.^5^ Implicit feedback systems, as used to manage COVID-19 in most jurisdictions, do not use an integrator (*K*_*i*_) but respond directly to measures such as case counts. As we shall see below, this results in less effective control.

When engaging a feedback controller into a situation where the system has previously been running in open-loop we need to manage the initial transient behavior to mitigate any large undesirable jumps in *v*(*t*). Generally there are techniques known as “bumpless transfer” that are employed in industry for this purpose (see for example [11]). The parameter *t*_0_ in (6) should be set to the time when the controller is engaged to initialize the control action and not for example backdated to the start of the data set, which could result in the cumulative error in the second term being quite large.

The constants *K*_*p*_ and *K*_*i*_ are tuning parameters through which the policy’s aggressiveness and robustness are designed. Using the SIMC formula from [7] described in the appendix, these are calculated to be *K*_*p*_ = 0.081, *K*_*i*_ = 4.4 · 10^−4^ for feedback with 14-day delay (2), and *K*_*p*_ = 0.20, *K*_*i*_ = 2.6 · 10^−3^ for the feedback with 2-day delay (3).

In the following section various simulations will be performed with the closed-loop system formed by the control law (6) and the full nonlinear SEEIQR model (7).

The explicit use of feedback does not require any particular policy update frequency. Frequent changes in public health policy may be undesirable in practice, leading policy makers to prefer bi-weekly or monthly updates rather than daily ones. As shown in the example below, this discretization is straightforward, and there is little loss in effective control as long as the policy actions are not too delayed. The interested reader is referred to references such as [12] for details.

## 3. Results

With the closed-loop system defined above, we examine the impact of various real-world situations - uncertainty in the model, the role of time delays in case counts, the impact of waning public compliance, and the introduction of changes including new variants and vaccinations.

### 3.1. Model Uncertainty

The effect of feedback on a highly uncertain pandemic is illustrated with Monte-Carlo simulations. Uncertainty in the parameter values is introduced by drawing parameters in the SEEIQR model from a range of values (see Appendix A) and simulating the dynamics, repeating this process for 400 realizations. Feedback control was based on hospital measured infections (2) with an uncertain delay from infection to hospitalization *d* (mean of 14 days, range: 4 - 28 days), and *R*_0_ varied between 1.04 and 5.18 (see Appendix A for variability introduced in other parameters). The controller used the tuning parameters *K*_*p*_ = 0.081, *K*_*i*_ = 4.4 · 10^−4^ in (6) (see Appendix B) and targeted a hospital-measured infected case count of 20 patients (approximately corresponding to a number of infections of 274). Figure 3 illustrates that the feedback (6) has effectively reduced the large variability in case counts and transferred it to different levels of interventions. In this simulation, we assumed that policy changes could be made only once every two weeks, discretizing the controller (6). We find that the feedback mechanism controls the pandemic despite the model uncertainty and broad range of time delays explored (from 4 to 28 days), in contrast to results presented in [13]. This robustness is due to the near-linear form of the dynamics of the pandemic when measured on a logarithmic scale (Appendix B and C).

**Figure 2.**
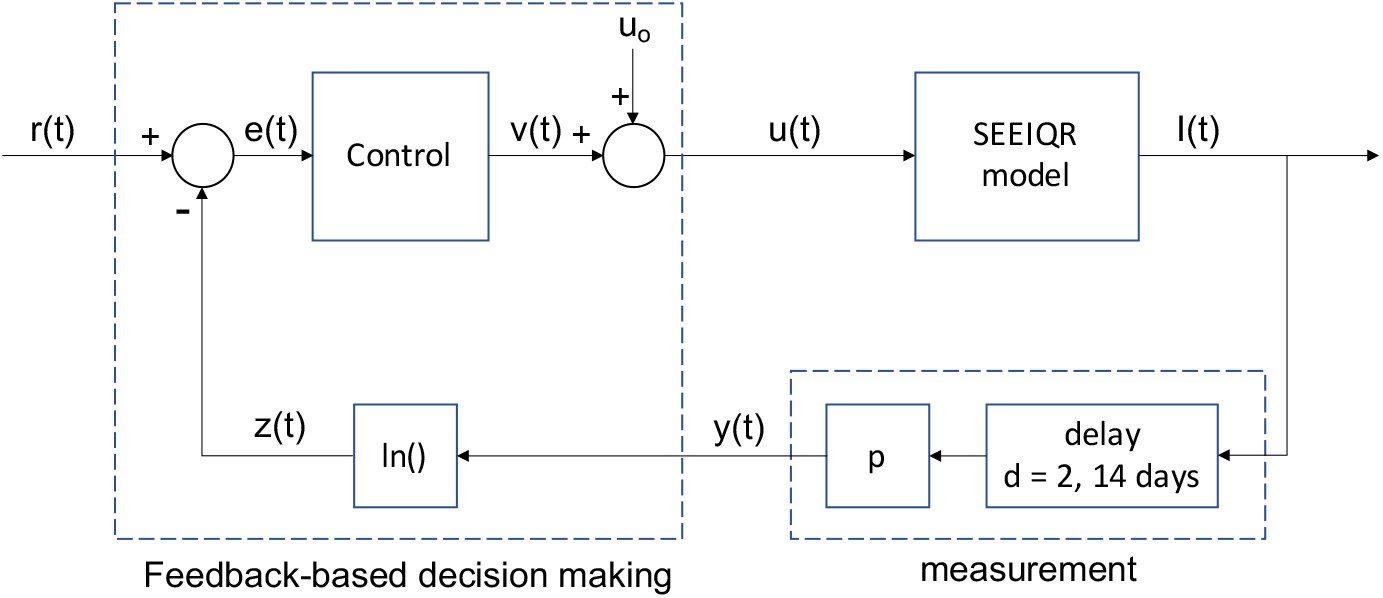
This block diagram illustrates the feedback configuration of the SEEIQR model (7), measurements of case counts (1), the control (6), and all transformations described in Section 2.

**Figure 3.**
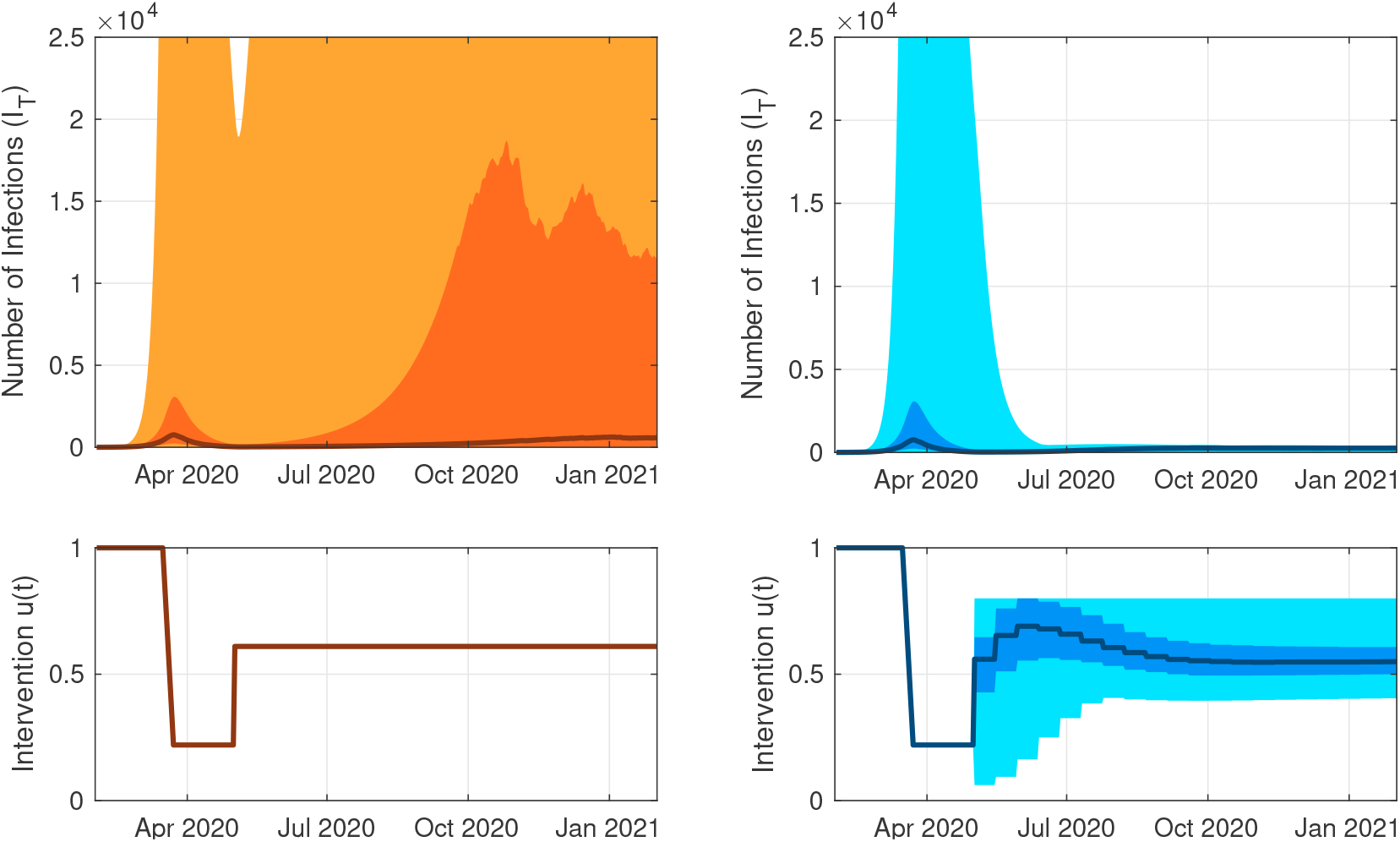
Accurate predictions are not required to design an effective policy to control the pandemic. Monte Carlo simulation results: The median is indicated with a thick line, the 25-75th percentiles and the min-max are indicated with shaded regions. Left: Under an open-loop policy , 40.25 % of the 400 realizations reach a maximal number of infections over 13500 after June 30th. Right: With the same dynamic feedback policy in place for all realizations, and bi-weekly intervention updates, 100 % of realizations maintain infections below 550 after June 30th. The variability in model parameters leads to adjustments in the interventions, which compensate for the differences between the models.

### 3.2. The Impact of Delays in Measurement

The response to an outbreak is evaluated and compared in two scenarios: with longer versus shorter delays. In Scenario 1 (equation (2)), feedback is based on counts from hospital testing, with a 14-day delay (controller parameters in (6): *K*_*p*_ = 0.081, *K*_*i*_ = 4.4 · 10^−4^, see Appendix B). In Scenario 2 (equation (3)), where feedback is based on counts from rapid public testing, with a 2-day delay (controller parameters in (6): *K*_*p*_ = 0.20, *K*_*i*_ = 2.6 · 10^−3^). The outbreak is initialized by 100 exposed individuals on day 0. As illustrated in Figure 4, reducing the delays in measuring the disease burden from 14 days down to 2 days has the overall effect of reducing both the cumulative case count by 80% and the average level of interventions *v*(*t*) by 6.7%. Time delay is well-known to place fundamental (i.e., not addressable via controller tuning) limitations on the performance of a closed-loop system. Thus, to the extent possible, reducing or eliminating time delays will improve the performance of the pandemic response.

**Figure 4.**
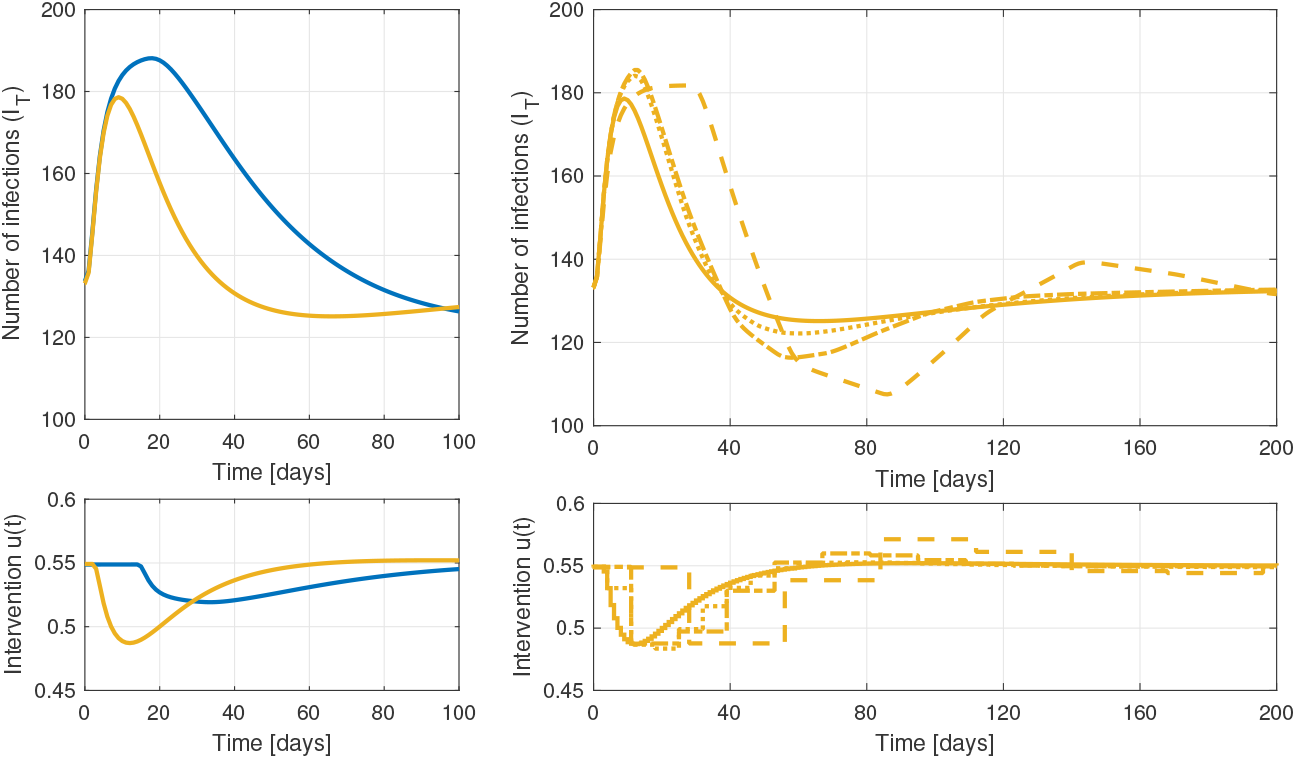
Minimizing delays in feedback improves performance: Left: Delays increase the time to detection of outbreaks and limit how aggressive policy responses can be. The top figure shows the response to an outbreak on day 0 (100 exposed individuals). Under feedback with 14 days of delay (blue), the case load as a result of the outbreak is ∫ *I*_*T*_ (*t*)*dt* = 2022, compared to ∫ *I*_*T*_ (*t*)*dt* = 406 for feedback with 2 days of delay (yellow). The bottom figure shows the corresponding interventions. Reducing the delay also reduces the cumulative interventions required to control the outbreak by 6.7 %. Right: The update frequency for interventions needs to be appropriate for the policy design and objectives. The behaviour of the feedback policy for 2 days of delay is similar when implemented with daily updates (yellow), weekly updates (dotted) or updates every two weeks (dash-dotted). Updates every four weeks (dashed) introduce too much delay and cause oscillations.

Discretization of these policy updates can be implemented with little loss in effective control, as long as the delay in policy updates is not too long with respect to the design objective (see Figure 4).

### 3.3. Varying Public Compliance

The effectiveness of non-pharmaceutical interventions has been hard to predict and may vary over time. Indeed, a key benefit of using feedback control is that appropriate action can be designed without knowing the exact effectiveness of these interventions. We simulated the case where the effectiveness of interventions was reduced by 0.075 on day 50. Even though we did not adjust the control system in any way, pandemic control with feedback adjusts the interventions to return the case count to the target (orange curves), whether we allowed for a continuous range of potential interventions (left) or a finite set (right). This robustness to changing public behaviour requires the use of an integral controller (*K*_*i*_), which takes the cumulative error into account. If, however, policy decisions were based only on current counts (no integral control: *K*_*i*_ = 0, holding proportional control *K*_*p*_ constant at 0.20), cases do not return to the target (purple) and are more sensitive to delays in policy interventions (right).

### 3.4. New variants and vaccinations

The effect of new, more contagious, variants and vaccinations is illustrated in Figure 6, again using a controller that tracks cases (measurement scenario 2 with a 2-day delay). A variant that is 2.5 times more contagious is introduced 425 days into the pandemic, reflecting the arrival of the Delta variant in March 2021 in British Columbia, with *R*_0*v*_ = 7.5 [14, 15]. Vaccination, with 95% efficacy against transmission, is introduced according to the vaccination rates in British Columbia^6^, starting in the week of December 19 2020. Vaccinations up to July 10th 2021 were taken into account, with a final vaccination rate of 70.92% of the population. The susceptible population was reduced by setting the parameter *p*_*vac*_ equal to the rate of first dose vaccinations in BC, delayed by 21 days to account for the maturation of the immune system. Importantly, the performance of the controller in Figure 6 was achieved without any information about variants or vaccines, using measured case counts only. If such variant information is available earlier, the peak may be further reduced by augmenting the feedback using that information in a feedforward fashion, i.e., by taking action earlier such that interventions are tightened or loosened proactively in anticipation of the arrival of a variant or vaccine respectively.

## 4. Discussion

A well-designed feedback policy can help mitigate the impacts of unknown and unknowable sources of uncertainty during a pandemic, including uncertainty in the dynamics, changing public compliance to recommended interventions, uncertainty about parameter values for a new disease, uncertainty about which interventions will work and how well, and the appearance of new variants or vaccines. All of these impacts are at best only roughly understood when they occur. While accurate models are more helpful than inaccurate models, these simulations illustrate that feedback-based policies designed on simple linear first-order plus delay approximations can successfully control a much more sophisticated SEEIQR model of the COVID-19 pandemic over a wide range of realistic conditions. Simulations also show that those feedback policies essentially transfer the uncertainty and variability in the case load to adjustments in the interventions. In nontechnical terms, if the case load starts to increase (decrease) for any reason, then the policy tightens (loosens) interventions in order to maintain the case load at its target (setpoint). The uncertainty-mitigating properties of feedback are fundamental in control theory.

Throughout the world, disease impacts through case counts or hospital load have been used to adjust restrictions. As discussed here, such feedback loops, using current values only (*K*_*i*_ = 0), are less efficient at controlling a disease than integral methods that also account for recent trends (6). Integration allows information to accumulate that disease impacts are moving beyond a target, allowing earlier action and improved control (Figure 5).

**Figure 5.**
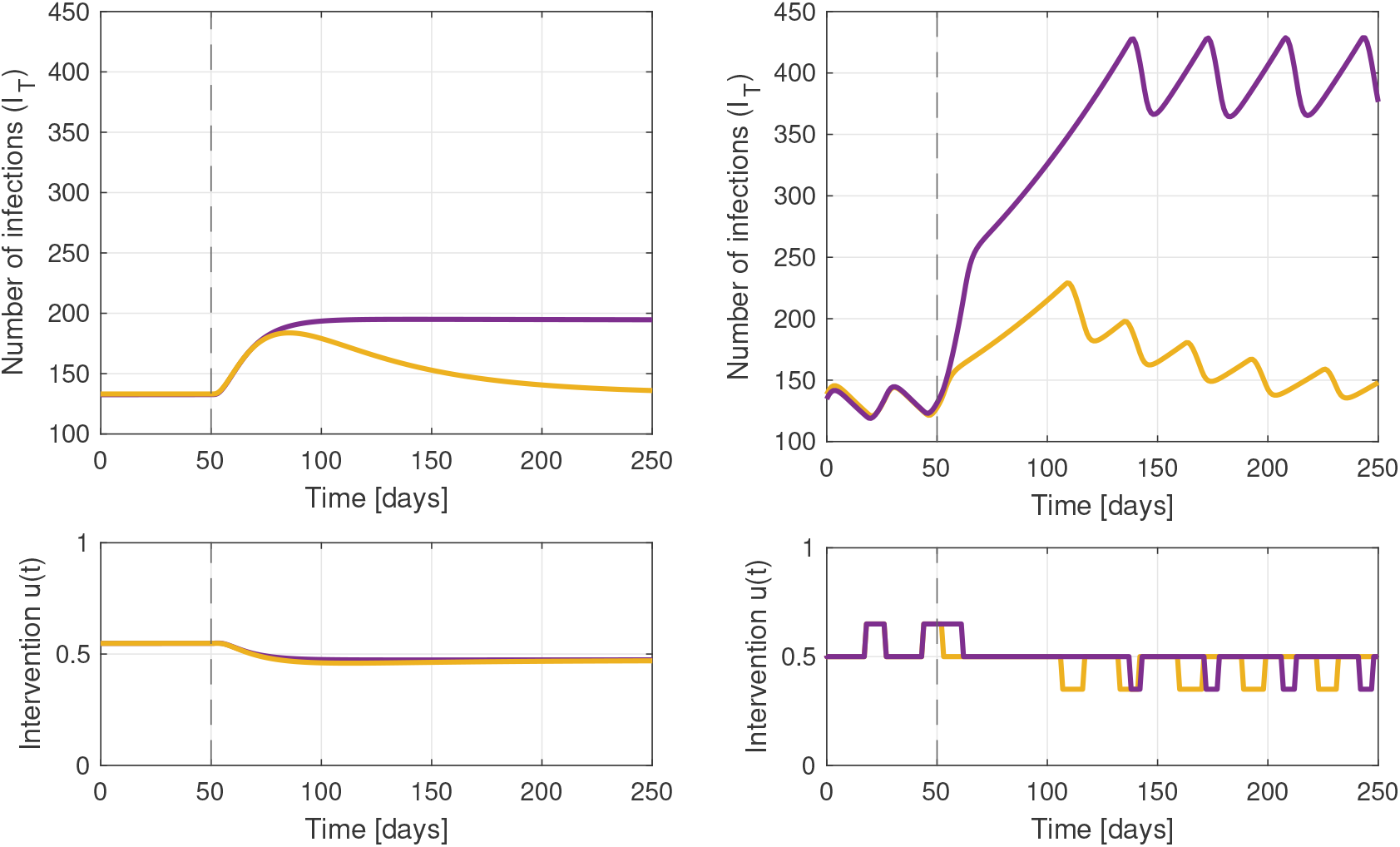
Feedback mitigates the effect of waning compliance to health directives. Left: After 50 days, the effectiveness of interventions decreases. Feedback control increases the level of interventions accordingly to stop pandemic growth. To fully counteract the (input) uncertainty, the cumulative error needs to be taken into account in the policy update (yellow; i.e., through the integrator term *K*_*i*_). If not (purple), the pandemic is stabilized at a sustained higher level of infections. Right: This effect is amplified if only a limited number of intervention levels are available. The area under the curve not using the cumulative error (purple) is 85 % higher than when the cumulative error is used (yellow).

With feedback control, any significant trend away from a target leads to a shift in policy, reducing variability in the quantity to be controlled (e.g., the case load). As a consequence, an effective policy can be designed even when the dynamics are uncertain; there is no need to wait for the development of a high-fidelity model. Of course, such models are useful to validate the performance of the feedback policy in simulations. Furthermore, any changes that cause a shift in the controlled variable (e.g., new variants, changing behaviour, vaccination) lead to automatic adjustments in the interventions. Such adjustments reduce the total burden of a disease, while also minimizing unnecessary restrictions.

As we have shown, delays in the gathering of information or in responding to that information limit the performance of a pandemic controller. Indeed time delays represent a fundamental limitation in closed-loop control systems [16]. An illustration of this problem is the recent landing on Mars of the Perseverance rover. The communication delay between Earth and Mars (7 min at the time of the landing), led NASA to coin the phrase “7 minutes of terror” as the delay made it impossible to control the descent from Earth. Time delays have to be explicitly included when designing feedback, lest the system becomes oscillatory or even unstable. In addition, as uncertainty in delays generally exists, robustness to delay variation has to be built in as well. Minimizing delays in the chain of measurement, reporting, decision making, and communication of restrictions is thus of critical importance. With smaller delays it is possible to reduce both the cumulative case counts as well as the cumulative severity of interventions. Rapid antigen testing and wastewater testing are examples of methods that can detect outbreaks early and so are important tools to consider in pandemic control.

For a new disease, an essential question is the translation of the control signal obtained by the controller into concrete, feasible public health measures, as there will initially be little known on how those map back to impact transmission (via *u*(*t*)). Input from epidemiologists, public health experts and behavioural scientists will be necessary to define and refine the array of measures that can be used to control a pandemic with increasing stringency.

This work used one of the simplest control algorithms available, the PID controller in its PI form as suggested in Stewart et al. [17]. Many other, more sophisticated control algorithms have been proposed since the COVID-19 pandemic started. One such class of control algorithms is the so-called model-based predictive control or MPC, (see e.g. [18]), which is one of the most studied control methods for this problem, see (e.g. [19, 20]). The theoretical advantage of MPC is that it can handle constraints, e.g., hard limits on variables such as interventions or the number of hospitalisations or ICU beds. However, in order to achieve that in practice, MPC requires accurate models and measurements. Another potential advantage of MPC is that, because it is a multivariable optimization-based control technique, the performance index to be minimized can be tailored to include not only public health indicators but socio-economic ones as well, thus allowing an adjustment of the compromise between managing the pandemic and preserving the economy. Nevertheless, for broader understanding and widespread application, the simple PI controller explored here brings many of the advantages of feedback control, without needing detailed and accurate knowledge of disease dynamics, which is often unknowable during a pandemic.

The control community was fast to realize that its tools could be used to help manage the pandemic, see e.g. [21] or [22]. Despite that, there has not been widespread use of feedback control theory by the public health and epidemiology community to guide decisions about when and how to enact public policies to manage the pandemic. This may be due to the fact that most of that work was published in control journals, typically not read by the public health community. Hence, the main objective of this article: reaching out to the public health community in order to raise awareness about control techniques and establish collaborations between the two communities. We hope to have achieved some initial progress toward this goal.

## Data Availability

All code will be available on GitHub

## Appendix A. SEEIQR model for COVID-19 in British Columbia

Simulation results are based on a two-group compartmental SEEIQR model, developed to estimate the effect of social distancing in early 2020 in British Columbia, Canada [5].

This model contains two groups, one representing individuals who are socially distancing and one representing individuals who cannot distance, for example due to their profession. The model compartments represent the number of susceptible (S), exposed (E1), pre-symptomatic and infectious (E2), symptomatic and infectious (I), quarantined (Q), recovered or deceased (R) for each group. The recovered (R) group does not affect infection rates and is omitted in this work.

The effect of vaccinations is included by reducing the size of the susceptible group. Variants with a different reproduction number have been included by additional compartments for each group, introducing a new pandemic drawing from the same susceptible population. The *S, E*_1_, *E*_2_, *I* and *Q* compartments for the distancing group are indicated with subscript *d*, compartments for the variant are indicated with subscript *v*. This does not account for possible re-infections. The effect of non-pharmaceutical interventions is introduced in this model through *u*(*t*), which can vary with time and affects the transmission from and to the distanced group. Public health policies are assumed to impact *u*(*t*) with *u*(*t*) = 0 representing complete restrictions resulting in no transmission involving the distanced group and *u*(*t*) = 1 representing no restrictions. For the original COVID-19 strain (with (t) omitted for readability), the equations for the group who are not physically distancing are:

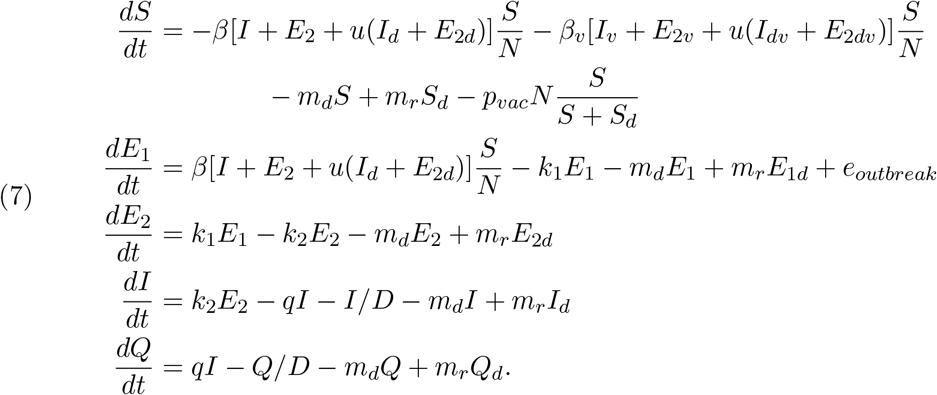

The transmission parameter, *β*, is related to the reproductive number, *R*_0_, according to 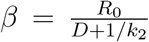 and 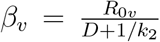 for the original strain and the variant respectively. *N* is the size of the population, *m*_*d*_ and *m*_*r*_ are the rates of movement to and from the physical distancing compartments. These parameters are assumed to be constant, with *m*_*d*_*/*(*m*_*r*_ + *m*_*d*_) of the cases initially in the distancing group. *p*_*vac*_ is the fraction of the population vaccinated per day. *k*_1_ is the rate of movement from the *E*_1_ to *E*_2_ compartment, *k*_2_ from the *E*_2_ to *I* compartment and *q* from the *I* to *Q* compartment. *D* is the mean duration of the infectious period. Outbreaks are simulated as imported cases (*e*_*outbreak*_) in the *E*_1_ compartment.

Analogous equations for the group that is distancing are:

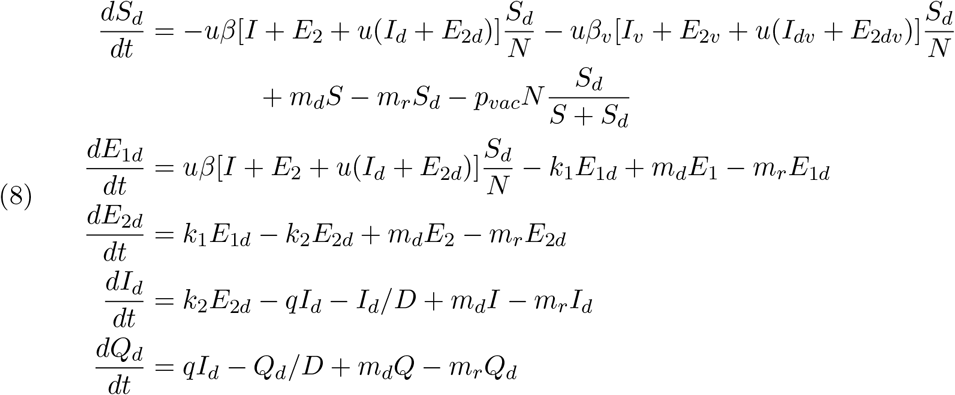

The equations for the new variant are driven by the introduction of one or more variant cases *e*_*variant*_(*t*) at a specified time *t*:

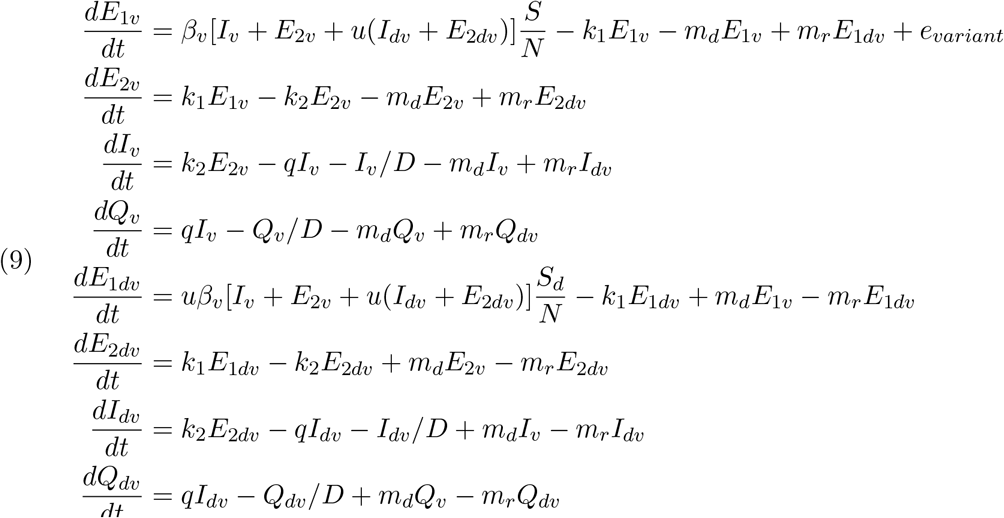

The nominal parameters used in the simulations are given in Table 1, as well as the range of parameters for the Monte-Carlo simulation presented in Section 3.1.

**Table 1.** Nominal model parameters and the range of values used to conduct Monte Carlo simulations. At the beginning of each of the 400 realizations, parameters were drawn from a normal distribution with average equal to the nominal values and standard deviations as indicated. The resulting models were used to simulate the pandemic with feedback control.

| | $D$ | $k_1$ | $k_2$ | $q$ | $R_0$ | Delay | $R_{0v}$ | $m_d$ | $m_r$ |
| --- | --- | --- | --- | --- | --- | --- | --- | --- | --- |
| Nominal | 5 | 0.2 | 1 | 0.05 | 3 | 14 | 7.5 | 0.1 | 0.02 |
| Standard deviation | 1 | 0.04 | 0.2 | 0.01 | 0.6 | 4 | NA | 0 | 0 |
| Minimum | 1.97 | 0.10 | 0.41 | 0.024 | 1.04 | 4 | NA | NA | NA |
| Maximum | 8.10 | 0.32 | 1.57 | 0.0784 | 5.18 | 28 | NA | NA | NA |

## Appendix B. Controller tuning: Simple internal model control (SIMC)

In this work, we use a simple proportional-integral (PI) feedback controller (6), tuned using SIMC (simple internal model control). PI and PID (proportional-integral-derivative) controllers are used in the majority of industrial control applications, and a multitude of tuning methods exist. The SIMC rules were developed for process control applications, and presented as “Probably the best simple PID tuning rules in the world” [7]. Process control applications often involve complex and nonlinear processes for which accurate models may not exist. The SIMC rules offer a practical solution for such problems, where a simple linear model is used to approximate a complex system and to identify appropriate parameters for feedback control. COVID-19 exhibits complex, nonlinear behaviour, yet can be approximated with a simple approximate model, and therefore the SIMC method can be used to tune a feedback controller.

The first step in SIMC is to find a linear first- or second-order plus time delay model that approximates the system dynamics. Section 2.2 describes the proposed model approximation, which is then given by:

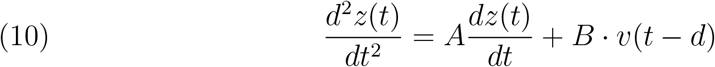

The parameter values, *A* = − 0.5787 and *B* = 0.1572, are derived using a least squares approximation to the nonlinear SEEIQR model (7) with nominal parameters in Table 1. These parameters are thus tuned to this pandemic model for British Columbia. The response of this approximate model to a step change in *v*(*t*) from *v*(*t*) = 0 to *v*(*t*) = 1, and its relation to the parameters *A, B* and *d*, is illustrated in Figure 7. The parameters *A, B* and *d* determine the time it takes for the system to respond (the delay *d*), the time constant to reach the constant slope, *τ*_2_ = −1*/A*, and the slope of the response, also referred to as the gain of the system, *κ* = −*B/A*.

**Figure 6.**
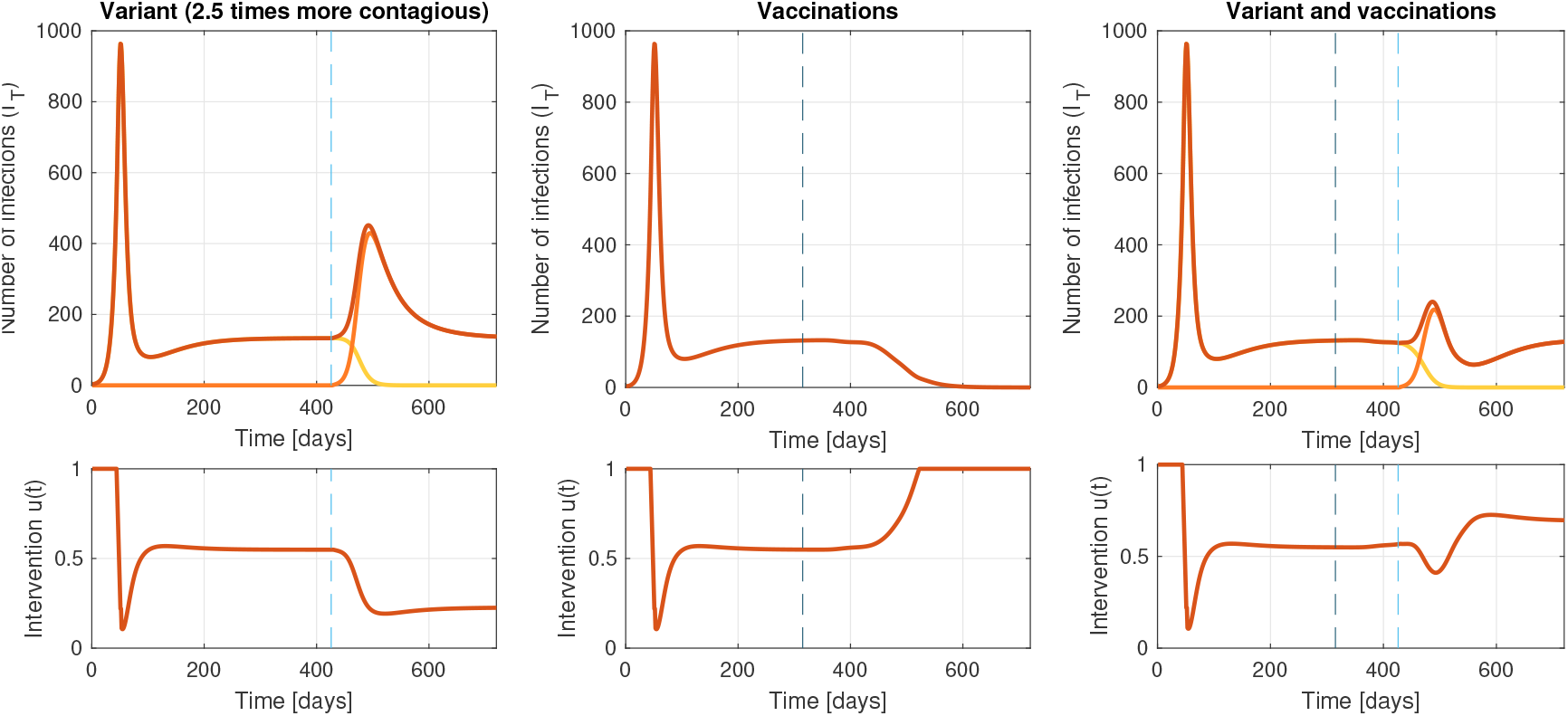
Feedback reduces the impact of the uncertainties inherent in a pandemic, such as the impact of vaccinations and new variants. Left: A new variant that is 2.5 times more infectious (*R*_0*v*_ = 7.5, introduced after 425 days indicated by a light blue dashed line) causes an initial increase in infections (dark orange line). Feedback control compensates with stricter measures that lead to extinction of the original strain (light line), while controlling the new strain at the target (orange line). Center: The effect of vaccine roll-out (start indicated by a dark blue dashed line) of *a priori* uncertain efficacy is mitigated by feedback, where restrictions are reduced in response to observed reduced case loads. Right: Feedback can mitigate the combined effect of new variants and vaccine roll-out. In this simulation, vaccination of 70.9% of the population with a vaccine that is 95% effective is insufficient to stop the more transmissible variant on its own. The feedback policy adjusts restrictions, and the number of infections remains controlled at the target.

**Figure 7.**
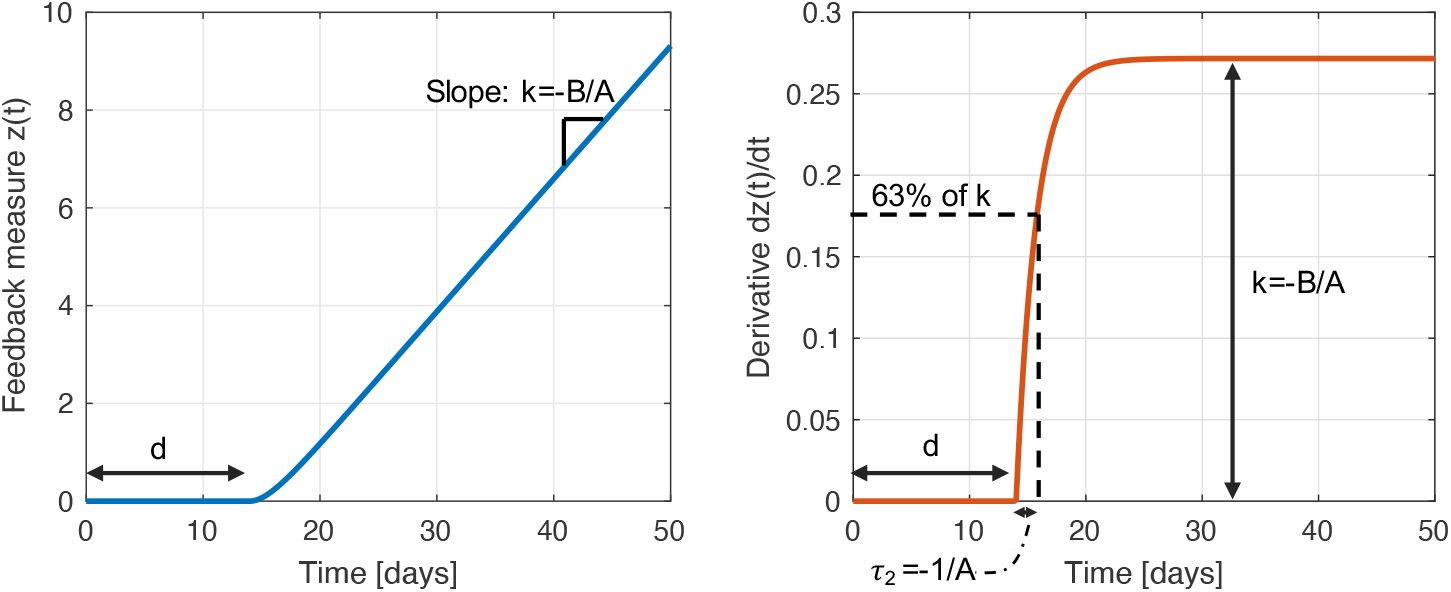
Left: Response of *z*(*t*) of approximate model to a step change in *v*(*t*) from *v*(*t*) = 0 to *v*(*t*) = 1. Right: Response of *dz*(*t*)*/dt* to a step change in *v*(*t*) from *v*(*t*) = 0 to *v*(*t*) = 1.

The second step in SIMC involves defining the controller (the function that maps *z*(*t*) to the intervention variable *v*(*t*) = *u*(*t*) − *u*_0_), with controller parameters determined by the approximate linear dynamics. For a system where the time delay *d* is larger than the time constant *τ*_2_, the SIMC rules [7] recommend a proportional-integral control policy (6), repeated here for readability.

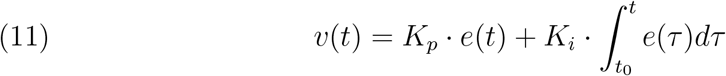

The controller parameters can then be calculated according to:

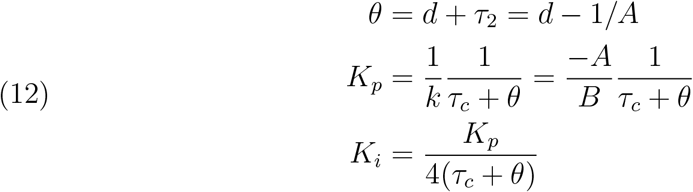

The SIMC tuning rules introduce a single tuning parameter *τ*_*c*_ that defines the aggressiveness of the controller, by which we mean that a small value of *τ*_*c*_ corresponds to an aggressive controller with a fast speed of response, while a large value of *τ*_*c*_ corresponds to a more conservative controller with a slower response, better stability and increased robustness to uncertainty in the model. In this work, we found that *τ*_*c*_ = 30 days for feedback with a long delay (*d* = 14 in Scenario 1) and *τ*_*c*_ = 15 for a 2-day delay (*d* = 2 in Scenario 2), represented a reasonable design tradeoff.

## Appendix C. Analytical results: SIR model

The linearization described in Section 2.2 is an approximation of the dynamics of the nonlinear SEEIQR model (Appendix A). For the special case of a simpler epidemiological model with an SIR structure with only one group (i.e. all individuals are socially distancing), this linearization is exact.

The SIR model, including social distancing measures, can be described as:

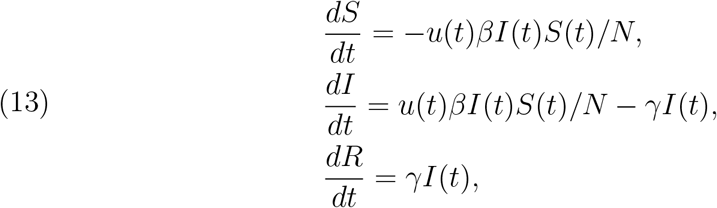

where *S*(*t*), *I*(*t*) and *R*(*t*) are the susceptible, infected and recovered states respectively, *N* the total population, *β* the transmission parameter and *γ* the recovery rate. The effect of social distancing is introduced as a factor *u*(*t*) that affects transmission, i.e. transmission at time *t* equals *β*(*t*) = *u*(*t*)*β*, with *u* = 1 corresponding to no distancing and *u* = 0 corresponding to reduction of transmission to 0. For simplicity of notation, we assume the measured variable *y*_*c*_(*t*) = *p* · *I*(*t*).

Without social distancing, this nonlinear SIR model can be linearized by assuming a constant susceptible population (*S*(*t*) = *S*_0_), or normalized constant susceptible population (*S*_0_*/N* = 1). The normalized infected population is then given by:

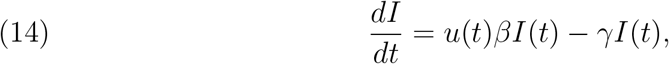

The input-output relation between interventions *u*(*t*) and *y*_*c*_(*t*) = *p* · *I*(*t*) remains nonlinear.

Assuming that a constant fraction *p* of the active infections is detected, the transformed variable *z*(*t*) = ln(*pI*(*t*)) is:

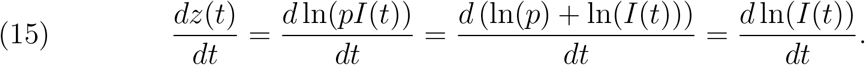

Equation 15 demonstrates why feedback is insensitive to the proportion, *p*, of detected cases. This result does not depend on the model structure (SIR or SEEIQR) and is essential for decision making and for updating *v*(*t*) and *u*(*t*) appropriately, given incomplete information. It does not matter what proportion of cases is captured in the feedback, and it is not even necessary to know what the captured proportion is, because only proportional changes in the measured variable matter.

The proposed log transformation for the SIR model, *z*_*c*_(*t*) = ln(*y*_*c*_(*t*)), and affine transformation *v*(*t*) = *u*(*t*) *− u*_0_ linearize the input-output model:

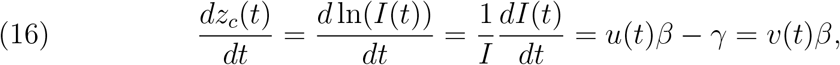

with *u*_0_ = *γ/β*. This result is now insensitive to scaling.

For this (one group) SIR model, the linearization is exact and the linearized model corresponds to a first order model. In a more complex epidemiological model such as the SEEIQR model, the linearization is approximate due to the exposed compartment, which also introduces additional dynamics that can be linearized by a lag as described in Section 2.2^7^. The SEEIQR model considered in this study contains two groups, which introduces a second source of non-linearity through the factor *u*(*t*)^2^. The simulation experiment included a series of step changes to the intervention *u*(*t*) to ensure it covers a representative range of responses to which we fit the linear model.

When solving the differential equation described by (16), it follows that the number of infections at any time *t*_*c*_ is a function of the initial case count and the cumulative (integral of) interventions up to time *t*_*c*_. Let *L*_0_ represent ln(*I*_0_), with *I*_0_ the number of infected individuals at time *t*_0_. The number of infected individuals at time *t*_*c*_ is then given by:

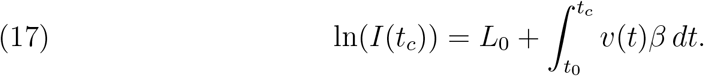

## Footnotes

1 Le Bourgeois Gentilhomme, Molière, 1670

2 https://health-infobase.canada.ca/src/data/covidLive/Epidemiological-summary-of-COVID-19-cases-in-Canada-Canada.ca.pdf accessed on Sept 14 2021

3 For the special case of a simple one-group SIR model the transformation leads to an exactly linear relationship, see Appendix C.

4 In practice, the fraction *p* of infections that are detected is often not known. However, as long as this fraction *p* is (piecewise) constant, it does not affect the linearized model (due to the log transformation above), and the scaling does not need to be known to implement effective control strategies (see Appendix C for details).

5 As noted above the true infection rate and the tested infection rate will differ by the false positive (FP) and false negative (FN) rates of the test used. So technically the closed-loop will converge to the desired rate of *measured* positives, which will differ from the true rate by an offset *c* = (*FP* − *FN*). These measurements may be combined in a cascaded control loop [10] where the inner control loop acts quickly on the fast measurements, while an outer control loop operates from the slower, accurate measurements and provides updates to the setpoint of the inner loop, correcting for the offset - in this way leveraging the strengths of different measurements.

6 https://health-infobase.canada.ca/covid-19/vaccination-coverage/, accessed on July 21 2021

7 Equation (5) is equivalent to 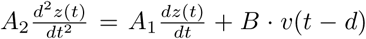 with *A*_2_ = 1 and *A*_1_ = *A*, while (16) corresponds to *A*_2_ = 0 and *A*_1_ = −1 and *d* = 0.

